# Quantification of stress and well-being using pulse, speech, and electrodermal data: Study concept and design

**DOI:** 10.1101/2020.05.01.20082610

**Authors:** Keisuke Izumi, Kazumichi Minato, Kiko Shiga, Tatsuki Sugio, Sayaka Hanashiro, Kelley Cortright, Shun Kudo, Takanori Fujita, Mitsuhiro Sado, Takashi Maeno, Toru Takebayashi, Masaru Mimura, Taishiro Kishimoto

**Author notes:** **Corresponding author:** Taishiro Kishimoto, MD, PhD, Department of Neuropsychiatry, Keio University School of Medicine, 35 Shinanomachi, Shinjuku-ku, Tokyo, 160-8582, Japan.

## Abstract

**Introduction:** Mental disorders are a leading cause of disability worldwide and, among mental disorders, major depressive disorder was highly ranked in years lived with disability. Depression has a significant impact in the field of occupational health because it is particularly prevalent during working age. On the other hand, there are a growing number of studies on the relationship between “well-being” and employee productivity. To promote healthy and productive workplaces, this study aims to develop a technique to quantify stress and well-being in a way that does not disturb the workplace.

**Methods and analysis:** This is a single-arm prospective observational study. The target population is adult (>20 years old) workers at companies that often engage in desk work; specifically, a person who sits in front of a computer for at least half their work hours. The following data will be collected: a) participants’ background characteristics; b) participants’ biological data during the 4-week observation period using sensing devices such as a camera built into or connected to the computer (pulse wave data extracted from the facial video images), a microphone built into or connected to their work computer (voice data), and a wristband-type wearable device (electrodermal activity data, body motion data, and body temperature); c) stress, well-being, and depression rating scale assessment data (New Occupational Stress Questionnaire, Perceived Stress Scale, Satisfaction With Life Scale, Japanese version of Positive and Negative Affect Schedule, Japanese Flourishing Scale, Subjective Well-being / Ideal Happiness, and Japanese version of Patient Health Questionnaire-9). The analysis workflow is as follows: (1) primary analysis, comprised of using software to digitalize participants’ vital information; (2) secondary analysis, comprised of examining the relationship between the quantified vital data from (1), stress, well-being, and depression; (3) tertiary analysis, comprised of generating machine learning algorithms to estimate stress, well-being, and degree of depression in relation to each set of vital data as well as multimodal vital data.

**Ethics and dissemination:** Collected data and study results will be disseminated widely through conference presentations, journal publications, and/or mass media. The summarized results of our overall analysis will be supplied to participants.

**Registration:** UMIN000036814

*STRENGTHS AND LIMITATIONS OF THIS STUDY:*

- This study evaluates stress and well-being using biomarkers such as heart rate, acoustic characteristics, and electrodermal activity.
- This study measures biomarkers over a long-term four-week period.
- This study will lead to the development of a machine learning algorithm to determine people’s optimal levels of stress and well-being.
- This is a government-funded study in which many different companies and institutions collaborated for a common purpose.
- There is a possibility that the algorithm’s prediction accuracy level may decrease when it is applied to demographic groups other than those studied here.

## INTRODUCTION

Mental disorders are a leading cause of disability worldwide and, among mental disorders, major depressive disorder was ranked number one in years lived with disability in 2017[1]. The lifetime prevalence of depression in Japan is estimated at 6.2% (2002–2006 estimate), which makes it the country’s most common mental illness[2]. The disease costs are also enormous and are estimated to exceed 3.09 trillion JPY (approximately 30 billion USD) per year[3]. Depression has a significant impact in the field of occupational health because it is particularly prevalent during working age (20–65 years of age). It is estimated that more than half of the social loss due to depression is attributed to loss of labor productivity through absenteeism and presenteeism[2]. The Japanese government has implemented measures against long working hours (Standards on limits of overtime work in 1998; the revision of the Industrial Safety and Health Act in 2006) and has introduced the stress check system (enforced as of December 2015), but there has not been a significant impact from those measures. In fact, the number of workers’ compensation claims related to mental disorders are increasing each year.

On the other hand, there are a growing number of studies on the relationship between “well-being” and employee productivity. The happiness of employees has been reported to be associated with creativity and productivity[4]. As the birthrate continues to decline and the aging population continues to increase in Japan, the working age population is also decreasing, requiring each employee to make the most of his/her abilities. Therefore, preventing negative factors such as depression and promoting well-being are major challenges for health management in the workplace and ensuring a stable economy.

Conventionally, it is known that heart rate variability (HRV) reflects autonomic nerve activity and serves as an index of psychological and physical stress. There are many suggested indicators for stress, including NN interval standard deviation (SDNN) and the percentage of successive RR intervals that differ by >50ms (pNN50) with time domain variables and low frequency (LF), high frequency (HF), and their ratio (LF/HF) as frequency domain variables[5]. In addition, techniques for estimating emotions and depressive symptoms have been developed based on the analysis of vocal formant frequencies[6]. Furthermore, electrodermal activity measured by a wearable device can reflect the activity of eccrine sweat glands that are controlled only by sympathetic nerve activity, and is therefore expected to be a stress indicator[7]. Previous studies have used the above mentioned approaches to measure subjects’ degree of stress; however, this prior research comprises only feasibility studies that have simply verified device performance, and/or studies with only a few patients or healthy individuals[8,9].

Moreover, such approaches are only used to evaluate the so-called “short-term stress” of the study period, and are not necessarily reflective of the effects of medium- to long-term stress. In a workplace environment, it is expected that regardless of whether employees experience temporary stress, there should also be situations where people feel a sense of freedom and accomplishment when a task is completed or a problem overcome. To date, very few studies have revealed the relationship between vital data and mid- to long-term stress and well-being in the workplace[10].

This study, which is funded by the Japan Agency for Medical Research and Development (AMED), is an industry-academia collaborative research project that aims to develop new techniques for evaluating mid- to long-term stress and well-being using technologies that will not obstruct normal work environments. By doing so, we hope to promote healthy workplaces and, in the end, to prevent depression in the prime of life.

### Research objectives

The general aim of this study is to develop a technique to quantify stress and well-being in a way that does not disturb the workplace. Our specific objectives are: (1) To evaluate the relationship between the obtained questionnaire-based stress and well-being scores and the employees’ vital data, which are collected using: a technique for extracting pulse waves from an image captured by a camera attached to the employee’s computer, a technique for extracting emotional components from speech, and a technique for measuring electrodermal activity using a wristband-type wearable device; and (2) to gather information regarding how and when employees are coping to reduce stress or promote enhanced well-being by comparing questionnaire-based stress and well-being scores and the employees’ vital data.

## METHODS AND ANALYSIS

### Study design

This is a single-arm prospective observational study.

### Participant criteria

#### Inclusion criteria

Adult (>20 years old) workers at companies that often engage in desk work; specifically, a person who sits in front of a computer for at least half their work hours (3.5 hours a day or more).

#### Exclusion criteria

People who correspond to any of the following groups are excluded from this study:

1. People currently receiving treatment for mental illness, such as depression;
2. People who suffer from diseases that may affect the acquisition of biometric information. For example, those who have a disease or disorder that affects pulse wave data measurement (persons who have paralysis or involuntary movements on their faces, or heart disease), those who have a disease or disorder that affects speech data measurement (speech difficulty caused by vocal cord extraction, etc.), or those who have a disease or disorder that affects measurement with wearable devices (persons with paralysis of the extremities or involuntary movement, etc.);
3. People who have difficulty operating a computer, such as using email or the internet;
4. People who cannot offer biometric information to researchers due to business/security reasons.

### Patient and public involvement

This study was supported by AMED at the stage of developing proof of concept for quantification of stress and well-being using pulse, speech, and electrodermal data. The study design was made by industrial doctors who served as consultants for some of the companies for which the participants of this study work. These industrial doctors, who are members of our research team, conducted preliminary meetings with the participants, and based on those meetings, they arrived at the question this study hopes to answer: whether stress and well-being can be quantified by pulse, speech, and electrodermal data. The results of this study will be made available to participants through debriefing sessions at each participating company.

### Data collection

Data will be collected according to the observation period schedule in Table 1.

**Table 1.** Schedule for data collection and evaluations during the study’s observation period

| Data collection |  | At the beginning | At the mid-point<br>(2 weeks) | At the end<br>(4 weeks) |
| --- | --- | --- | --- | --- |
| A) Collection of background factors | Background characteristics (sex, age, department, work content, duration of service, etc.)<br>Past stress check data, etc. | ✓ | If participant's environment changes, data will be updated. |  |
| B) Collection of data using external sensors | Pulse wave, speech, electrodermal activity, etc. | Acquired during business hours |  |  |
| C) Stress, well-being, and depression assessment using rating scale; self-reported daily condition | New Occupational Stress Simple Questionnaire (revised version): Estimated completion time, 5 minutes | ✓ | If participant's environment changes, data will be updated. |  |
|  | Perceived Stress Scale (PSS): Estimated completion time, 1 minute |  |  | ✓ |
|  | Satisfaction With Life Scale (SWLS): Estimated completion time, 1 minute |  |  | ✓ |
|  | Japanese version of Positive and Negative Affect Schedule (PANAS): Estimated completion time, 1 minute |  | ✓ | ✓ |
|  | Japanese version of Flourishing Scale (FS-J): Estimated completion time, 1 minute |  |  | ✓ |
|  | Subjective Well-being / Ideal Happiness: Estimated completion time, 1 minute |  | ✓ |  |
|  | Japanese version of Patient Health Questionnaire-9 (PHQ-9): Estimated completion time, 1 minute |  | ✓ | ✓ |
|  | Self-reported daily condition: Estimated completion time, 1 minute | Daily at the end of work (not required) |  |  |

#### A) Collection of background factors

After obtaining written consent, the following information will be obtained from each participant:

1. Sex, age, job department, job content, duration of service, position/title, family composition, work commute, household income, etc.;
2. Information on past medical checkups and stress check information (with consent of participant);
3. Any current illnesses and prescriptions.

#### B) Collection of biological data with sensing devices

Biological information will be recorded at participants’ workplaces during the 4-week observation period using methods B-1 through B-3, as described below:

##### B-1) Pulse wave data

Participants install software on their work computers that uses a camera built into or connected to the computer to record video images of the participant; the pulse wave data is extracted from the facial video images. The pulse wave data is automatically sent to cloud storage through the software. Participants are asked to start the software when they arrive for work; the software must also be restarted if the participant’s computer is put into sleep mode. This contactless pulse wave sensing system has a strong correlation in the R-R interval values compared to data obtained using ECG (r2= 0.978, p < 0.00001)[11].

##### B-2) Voice data

Participants install software on their work computers that uses a microphone built into or connected to their work computer to record the emotional components (pitch, speed, etc.) of participants’ speech data. The emotional component data is automatically sent to cloud storage through the software. Participants are asked to start the software when they arrive for work; the software must also be restarted if the participant’s computer is put into sleep mode.

##### B-3) Electrodermal activity data, body motion data, and body temperature

Participants are asked to wear the Embrace2 wristband-type wearable device, made by Empatica, Inc., continuously during work hours. The device is equipped with an electrodermameter, accelerometer, gyroscope, and thermometer[12,13].

C) Collection of stress, well-being, and depression rating scale assessment data; self-reported daily condition Researchers will send participants an email with a unique URL link for a unique website where participants can answer questionnaires on stress and well-being online. The evaluation scales and their estimated completion times are as follows (see Table 1 for the evaluation schedule):

- New Occupational Stress Questionnaire (modified version)[14]
- Perceived Stress Scale (PSS)[15]
- Satisfaction With Life Scale (SWLS)[16]
- Japanese version of Positive and Negative Affect Schedule (PANAS)[17]
- Japanese Flourishing Scale (FS-J)[18]
- Subjective Well-being / Ideal Happiness[19,20]
- Japanese version of Patient Health Questionnaire-9 (PHQ-9)[21]
- Self-reported daily condition: an email with a unique URL will be sent to participants every business day; participants will input their condition (stress level, emotions, etc.] and sleep quality for the day, and include any special notes in 1–2 sentences.

### Data analysis

The analysis workflow is as follows: (1) primary analysis, comprised of using software to digitalize participants’ vital information; (2) secondary analysis, comprised of examining the relationship between the quantified vital data from (1), stress, well-being, and depression; (3) tertiary analysis, comprised of generating machine learning algorithms to estimate stress, well-being, and degree of depression in relation to each set of vital data as well as multimodal vital data. The primary analysis is conducted with technology already established by Panasonic and NEC, who are industrial collaborators in this study. In this research, the results of the primary analysis are used to generate machine learning algorithms for the secondary and tertiary analyses.

#### Primary analysis

##### Primary analysis of pulse wave data

Software from Panasonic installed on participants’ work computers uses a camera to capture facial images of participants using facial detection. Based on skin color changes from blood flow in the images, the software extracts pulse wave data for the participant. The pulse wave data will then be clarified using filtering and noise removal techniques. SDNN, root mean square of successive differences in RR intervals (RMSSD), Lorentz plot (L / T value), LH/HF ratio, and Tone-Entropy are calculated from the facial video images (1/30 second unit).

##### Primary analysis of voice

Software from Panasonic installed on participants’ work computers uses a microphone to acquire speech data from participants. From this data, voice activity detection (VAD; presence/absence of voice), power (volume of speech), pitch, tension (strength of speech), and speech rate data are extracted (in units of 0.5 seconds), and an emotion estimate is calculated based on the results.

##### Primary analysis of electrodermal activity

The Embrace2 wristband-type wearable device from Empatica, Inc., is equipped with an electrodermameter, accelerometer, gyroscope, and thermometer, which are used to record and analyze electrodermal activity, acceleration, angular velocity, and skin temperature.

#### Secondary analysis

Each result from the primary analyses of pulse wave, speech, and electrodermal activity data will be compared with the self-assessment results for stress, well-being, and depression. Then, we will determine the relationships between the vital data, stress, well-being, and depression. For example, such an investigation could be done by comparing the stress and well-being score quartiles with the vital data, or comparing them among subjects grouped according to depression symptom severity.

#### Tertiary analysis

We will attempt to build a machine learning model to predict depression, stress, or well-being based on single modalities. Various methods, such as support vector machine, decision tree, and deep learning, are used for the machine learning analysis. Machine learning algorithms that estimate stress, well-being, and degree of depression are generated from each of these vital data sets. We will attempt to build a model not only for single modalities, but also one that can utilize all the modalities together.

### Sample size

Based on the data obtained from the pilot study (28 cases) conducted prior to this study, we estimated the PSS scores using machine learning analysis of pulse wave and speech data. A gradient boosting decision tree was used for the machine learning algorithm, and hyper parameter was adapted by random search. Accuracy verification is based on 3-fold cross validation. In order to examine the accuracy for each data size, an arbitrary number of data points were extracted at random, and the accuracy for the data size was calculated. The pilot study sample size of 28 subjects was divided and incremented for prediction. The error in the PSS score range of 0–40 corresponds to a score of 2 when an error in the predicted value of PSS of up to around 5% is warranted. When aiming for a root mean square error of 2 or less, we found that 200 cases are required, as shown in Figure 1.

**Figure 1.**
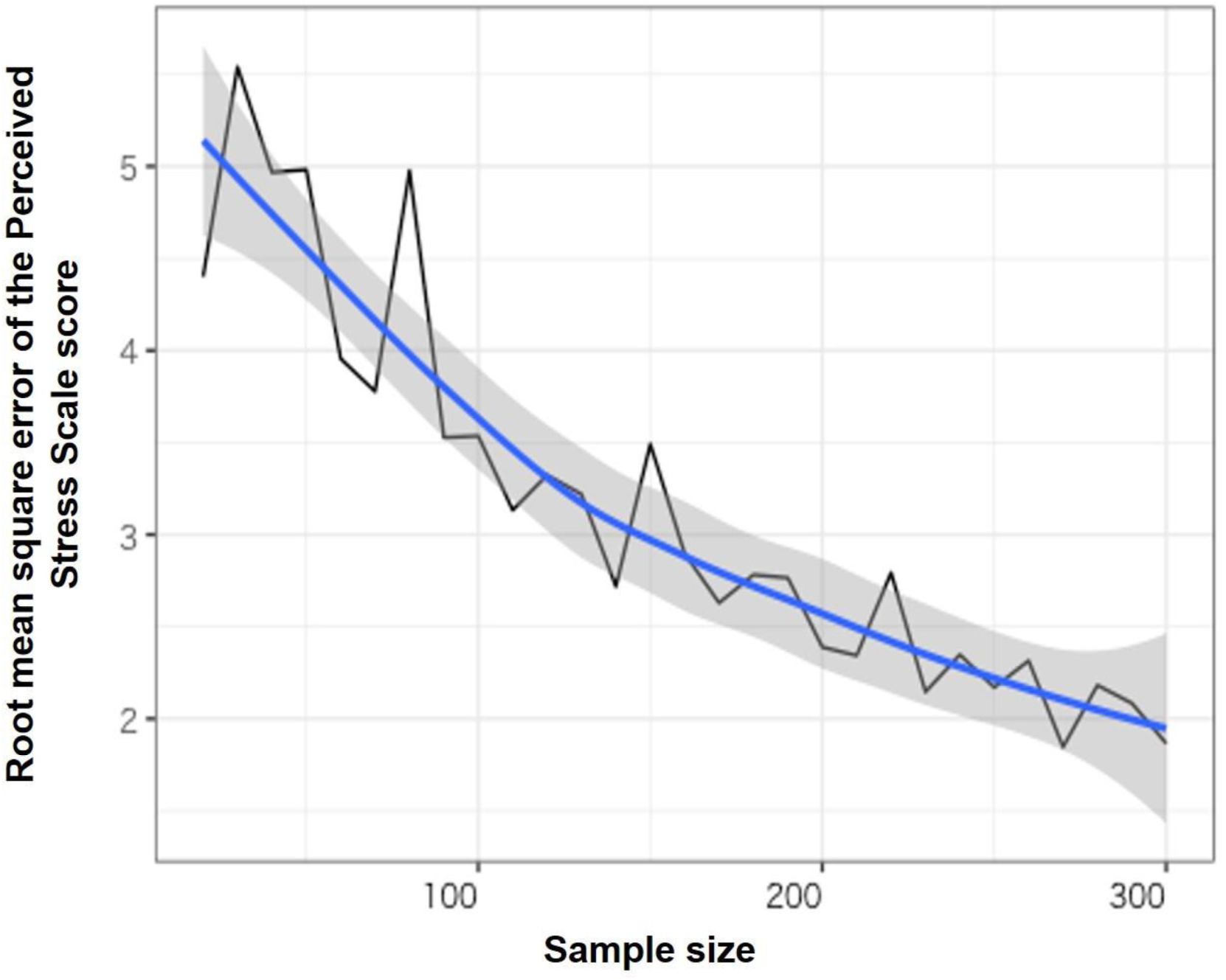
Sample size and root mean square error of the PSS score in the pilot study

## Data Availability

Not applicable

## ETHICS AND DISSEMINATION

### Ethical considerations

Our study has received approval from the institutional review board at Keio University School of Medicine. Approval was granted on April 22, 2019. This study is registered in the University Hospital Medical Information Network (UMIN) (UMIN000036814). Participants’ inclusion will be voluntary. Written consent will be obtained from every participant. Participants will be free to withdraw from the study at any time.

### Dissemination

Collected data and study results will be disseminated widely through conference presentations, journal publications, and/or mass media. The summarized results of our overall analysis will be supplied to participants.

## Acknowledgments

The authors are grateful to Mr. Yuichiro Suzuki, Mr. Yoshifumi Onishi, Ms. Momoko Kitazawa, Mr. Shunya Kurokawa, Mr. Michitaka Yoshimura, Mr. Kuo-ching Liang, Mr. Asuka Koshi, Ms. Yuki Ishikawa, Ms. Yoko Usami, Ms. Kumiko Hiza, and Ms. Hiromi Mikami for supporting data collection and management in this study.

## Authors’ contributions

KI, KM, and TK conceived the original study concept. KI designed and managed the study, and wrote the initial draft of the manuscript. KM and KS designed and managed the study. TK designed and supervised the study, and assisted in drafting the manuscript. All authors contributed to the design of the study, protocol development, its implementation, and critically reviewed the manuscript. All authors approved the final version of the manuscript, and agree to be responsible for the accuracy and integrity of the work.

## Funding statement

This work was supported by the Japan Agency for Medical Research and Development (AMED) (grant number 18le0110008h0001). The funding source did not participate in the design of this study and will not have any hand in the study’s execution, analyses, or submission of results. Japan Agency for Medical Research and Development (AMED) 20F Yomiuri Shimbun Bldg. 1-7-1 Otemachi, Chiyoda-ku, Tokyo 100-0004 Japan Tel: +81-3-6870-2200, Fax: +81-3-6870-2241

## Competing interests statement

All authors have no conflict of interest.

## Notes

### Competing Interest Statement

The authors have declared no competing interest.

### Clinical Trial

UMIN000036814

### Clinical Protocols

https://upload.umin.ac.jp/cgi-open-bin/ctr/ctr.cgi?function=brows&action=brows&recptno=R000041686&type=summary&language=J

